# No evidence for differential gene expression in major depressive disorder PBMCs, but robust evidence of elevated biological ageing

**DOI:** 10.1101/2020.09.04.20165340

**Authors:** John J. Cole, Alison McColl, Robin Shaw, Mary-Ellen Lynall, Philip J Cowen, Peter de Boer, Wayne C Drevets, Neil Harrison, Carmine Pariante, Linda Pointon, NIMA consortium, Carl Goodyear, Edward Bullmore, Jonathan Cavanagh

## Abstract

**Background:** The increasingly compelling data supporting the involvement of immunobiological mechanisms in Major Depressive Disorder (MDD) might provide some explanation of the variance in this heterogeneous condition. Peripheral blood measures of cytokines and chemokines constitute the bulk of evidence with consistent meta-analytic data implicating raised proinflammatory cytokines such as IL6, IL1β and TNF. Among the potential mechanisms linking immunobiological changes to affective neurobiology is the accelerated biological ageing seen in MDD, particularly via the senescence associated secretory phenotype (SASP). However, the cellular source of immunobiological markers remains unclear.

**Aims:** Pre-clinical evidence suggests a role for peripheral blood mononuclear cells (PBMC), thus here we aimed to explore the transcriptomic profile using RNA sequencing in PBMCs in a clinical sample of people with various levels of depression and treatment response comparing it with that in healthy controls (HCs).

**Method:** Transcriptomic analysis of peripheral blood mononuclear cells.

**Results:** The data showed no robust signal differentiating MDD and HCs. There was, however, significant evidence of elevated biological ageing in MDD vs HC.

**Conclusions:** Future work should endeavour to expand clinical sample sizes and reduce clinical heterogeneity. The exploration of RNA-seq signatures in other leukocyte populations and advances in RNA sequencing at the level of the single cell may help uncover more subtle differences. However, currently the subtlety of any PBMC signature mitigates against its convincing use as a diagnostic or predictive biomarker.

Highlights
- PBMCs showed no differential transcriptomic signature between depressed cases and healthy controls suggesting that the cellular source of the immune signature seen in depression may be from a different cell group.
- There was significant evidence of accelerated biological ageing in major depression compared to healthy controls.

## Introduction

Major Depressive Disorder (MDD) remains one of the most aetiologically opaque of human disorders, yet one that continues to exert a powerfully negative toll on human health – physical as well as mental. MDD is both heterogeneous in its phenotypic expression and complex is its genetic and physiological correlates. Among the latter there are increasingly compelling data supporting the involvement of immunobiology in MDD. However, the mechanisms underpinning this relationship remain unclear. Peripheral blood measures constitute the bulk of evidence with consistent meta-analytic data implicating raised proinflammatory cytokines. The most comprehensive genome-wide association study (GWAS) to date on MDD used 7 major cohorts and identified 44 independent loci and 153 genes^1^. Forty-five of these were in the extended major histocompatibility complex (MHC), which is central to acquired immunity and to leukocyte interactions.

Whole-*transcriptome* studies offer another variant of genome-wide search for disease-related mechanisms by measuring mRNA expression levels of each gene in a relevant tissue. RNA sequencing (RNA-seq) uses next-generation sequencing to provide a quantitation of RNA or gene expression. Recent studies have used this method in MDD. One of the largest examined a total sample of 922 people (463 with MDD and 459 health controls) and sequenced RNA from whole blood^2^. A relatively small number of genes were found to be associated with MDD (29) at a very relaxed false discovery rate (FDR) threshold of 0.25. With the more customary and restrictive FDR threshold of 0.05, no significant genes were found. They also showed modest enrichment for the IFN α/β pathway, which included three significant genes at FDR<0.25.

A number of potential mechanisms have linked immunobiological changes to affective neurobiology. Among these is the accelerated biological ageing seen in MDD. Immune cell senescence has a well-documented effect on both epigenome and transcriptome^3^. MDD has also been linked to the senescence associated secretory phenotype (SASP), a dynamic secretory molecular pathway indicative of cellular senescence^4^. This speaks to a more elaborate biology linking cell biology, transcriptome and inflammatory proteins produced by the cell.

The cellular source of immunobiological markers in depression remains a key unanswered question. PBMCs are a key source of peripheral cytokines and pre-clinical models have suggested some PBMC subsets can enter the brain and contribute to onset of sickness behaviour in the context of stress. McKim et al showed that IL1β-producing monocytes were recruited into the brain during stress and associated with increased levels of behavioural anxiety^5^. Menard et al confirmed McKim et al’s findings showing monocyte recruitment to vessels and ventricular space within the brain^6^. Recruited monocytes can release proinflammatory cytokines such as IL6 into parenchyma to act locally on neurons and glia. Garré et al used a viral model to show that defective cortical dendritic spine remodelling, and subsequent memory impairment were due to circulating CX3CR1^+^ monocyte-derived TNF^7^. Importantly, they showed that these exogenous monocytes, and not microglia, were necessary for these effects.

Given the weight of the preclinical evidence suggesting a role for PBMCs, we aimed to explore the transcriptomic profile using RNA-seq in PBMCs in a clinical sample of people with various levels of depression and treatment response and compare with that in healthy controls.

We aimed to answer the following research questions.

1. Is there evidence of differential gene expression between healthy controls and MDD or between healthy controls and sub-types of MDD?
2. Is there evidence of elevated immune ageing MDD compared to healthy controls?

## Methods

### Participants

This was a non-interventional study, conducted as part of the Wellcome Trust Consortium for Neuroimmunology of Mood Disorders and Alzheimer’s disease (NIMA). There were five clinical study centres in the UK: Brighton, Cambridge, Glasgow, King’s College London, and Oxford. All procedures were approved by an independent Research Ethics Committee (National Research Ethics Service East of England, Cambridge Central, UK; approval number 15/EE/0092) and the study was conducted according to the Declaration of Helsinki. All participants provided informed consent in writing and received £100 compensation for taking part.

### Sample and eligibility criteria

We recruited four groups of participants: treatment-resistant depression, treatment-responsive depression, untreated depression, and healthy volunteers. Eligibility criteria can be viewed in full in Supplementary Information.

Patients were assigned to one of three subgroups or strata, per protocol:

i. treatment-resistant (DEP+MED+) patients who had total Hamilton Depression Rating Scale (HAM-D) score > 13 and had been medicated with a monoaminergic drug at a therapeutic dose for at least six weeks;
ii. treatment-responsive (DEP-MED+) patients who had total HAM-D < 7 and had been medicated with a monoaminergic drug at a therapeutic dose for at least six weeks; and
iii. untreated (DEP+MED-) patients who had HAM-D > 17 and had not been medicated with an antidepressant drug for at least six weeks.

### Questionnaire assessments

Psychological symptoms and childhood adversity were assessed by administration of questionnaires as previously described^8^ (see Supplementary Information). Baseline depression severity was rated using the 17-item HAM-D.

### Sampling and isolation of PBMCs

Whole blood was collected in K2EDTA tubes (BD, USA) by peripheral venepuncture and allowed to cool to room temperature for a minimum of 45 minutes. PBMCs were collected from the interphase following density gradient centrifugation. RNA was extracted using the RNeasyMini Kit (Qiagen, Germany) as per the manufacturer’s instructions. RNA was eluted in 50ul RNase-free H_2_O and stored at −80°C before being sent for sequencing.

### RNA-sequencing and processing

PBMC samples were taken from four separate population groups as follows: **44** healthy controls, **94** MDD treatment-resistant, **47** MDD treatment-responsive, **46** MDD untreated patients. All PBMC samples had an RNA Integrity Number (RIN) ≥ 8 and were analysed for gene expression levels by RNA-Seq to an average depth of 54.5 million read pairs. Reads were trimmed using Cutadapt 1 (version cutadapt-1.9.dev2)^9^. The reference used for mapping was the Homo sapiens genome from Ensembl, assembly GRCh38, annotation version 84. Reads were aligned to the reference genome using STAR 2 (version 2.5.2b)^10^. Reads were assigned to features of type ‘exon’ in the input annotation grouped by gene_id in the reference genome using featureCounts 3 (version 1.5.1). Strandedness was set to ‘reverse’ and a minimum alignment quality of 10 was specified. After filtering for only protein coding genes, we observed a median of **40** million exonic aligned reads per sample (>85%).

### RNA-sequencing differential expression analysis

Differential expression analysis was performed using DESeq2 (version 1.18.1)^11^. The count matrix was initially filtered to include only coding genes, with a mean of > 1 read per sample. For the comparisons of binary clinical covariates (e.g. gender, tobacco) one group was compared to the other. For continuous clinical covariates (e.g. age, BMI) the patients in the lower quartile were compared to those in the upper quartile. No additional covariates were used in the DEseq2 model when comparing clinical covariates. For the comparisons between HC group and the MDD groups the 15 clinical covariates (**Figure 1b**) identified as having > 5 significant associated genes (adjusted p < 0.01) and “batch” were included as covariates in the model. To control for extreme outlier values typical in large and heterogeneous datasets, a Cooks cut-off of 0.2 was used. All other parameters were left to default. Significance was set at an adjusted p of < 0.01. For full details see the Supplementary Information.

**Figure 1.**
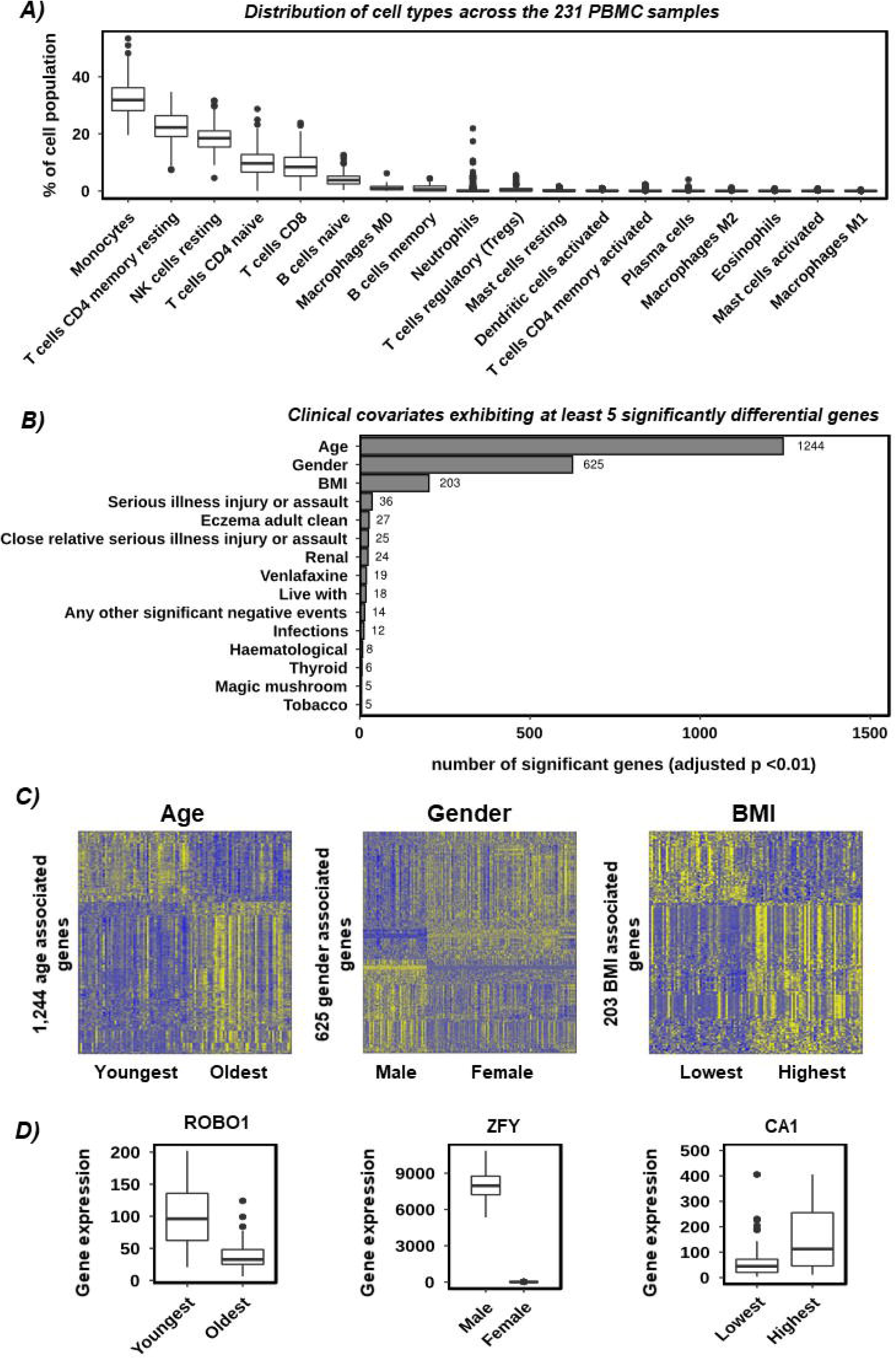
Quality control and identification of confounding variables. **A)** Distribution of immune cell types across all 231 PBMC samples. Cell types are shown on the x-axis, and the percentage of the cell population that is described by each cell type is shown on the y-axis. Each box represents all 231 samples. **B)** Bar chart showing the number of significantly different genes (DESeq2 adjusted p value < 0.01) across all clinical parameters with at least 5 significant genes. **C)** Gene expression heatmaps highlighting the size and consistency of the confounding effects of Age (left), Gender (middle), and BMI (right) on the PBMC RNAseq data. Samples are given by column and differentially expressed genes (adjusted p < 0.01) by row. Colour intensity indicated row scaled (z-score) gene expression, with blue as low and yellow as high. **D)** Gene expression boxplots of the most significantly different gene between youngest and oldest (ROBO1), male and female (ZFY), and lowest and highest BMI (CA1). Sample groups are shown on the x-axis and gene expression values (Corrected DESeq2 normalised counts) on the y-axis.

### Deconvolution analysis

The per sample distribution of cell types was estimated by Cibersort^12^, using the Deseq2 normalised expression values (no additional covariates) as the mixture file, and the LM22 (22 immune cell types) signature gene file. Quantile normalisation was disabled. All other parameters were left to default.

### RNA-sequencing randomised cases and controls

The 231 samples were randomised using the r function “sample” (without replacement), and were then split into two random groups, one with 44 samples and one with 187 samples (in line with the real group distribution and n). These two groups were then differentially compared using DESeq2 as described above. For full details see the Supplementary Information.

### Co-expression analysis

The co-expression network cluster analysis was based on the analysis performed by Le et al^15^ and used their code as a template. The method is detailed in full in Supplementary Information. Briefly, a correlation tree was generated from the expression matrix based on Pearson correlation coefficients and a topological overlap matrix. Clusters were identified by cutting the tree at a height of 0.95. To identify any clusters with significantly different gene expression between HC and MDD samples, a metagene for each cluster was generated using per gene Z-scores. For each cluster the mean expression z-score across all genes in that cluster was calculated, for each sample. The resultant scores for the HC samples were compared to that of the MDD samples using an unpaired, two-tailed T-test. p values were adjusted using the Benjamini-Hochberg procedure.

### Expression microarray analysis

The GSK-HiTDiP MDD^16^ microarray data was downloaded from GEO (GSE98793) and the 22 samples that were reported to have failed QC were removed. The expression data was then quantile normalised using Limma^17^. Unannotated probe sets were removed. To control for genes represented by several different probe sets, Jetset^18^ was used to select the probe set for each gene with the highest Jetset score. This resulted in 20,191 valid probe sets. Differential expression analysis was performed between the HC and MDD groups using Limma, and included batch, age, gender and anxiety as additional covariates. All other parameters were left to default. The quantile normalised expression values were corrected for batch using Limmas “removeBatchEffect” function.

### RNA-sequencing biological age meta-genes

A list of PBMC age associated genes was identified by using Deseq2 to compare the samples of lowest to highest quartile of age, as described above. Next the expression values (non-corrected but outlier capped) for the PBMC age related genes were scaled (per gene z-score), with the sign inversed for genes that were downregulated with age. Finally, the mean scaled value (across all sig genes) per sample was calculated. This value was considered as the samples biological age. The samples biological age was then plotted against the samples chronological age, and the spearman correlation value determined. To optimise this metric, we repeated over a range of adjusted p and log2fold change cut-offs and selected the combination with the greatest correlation with patient age. For full details see the Supplementary Information.

## Results

**Table 1.**
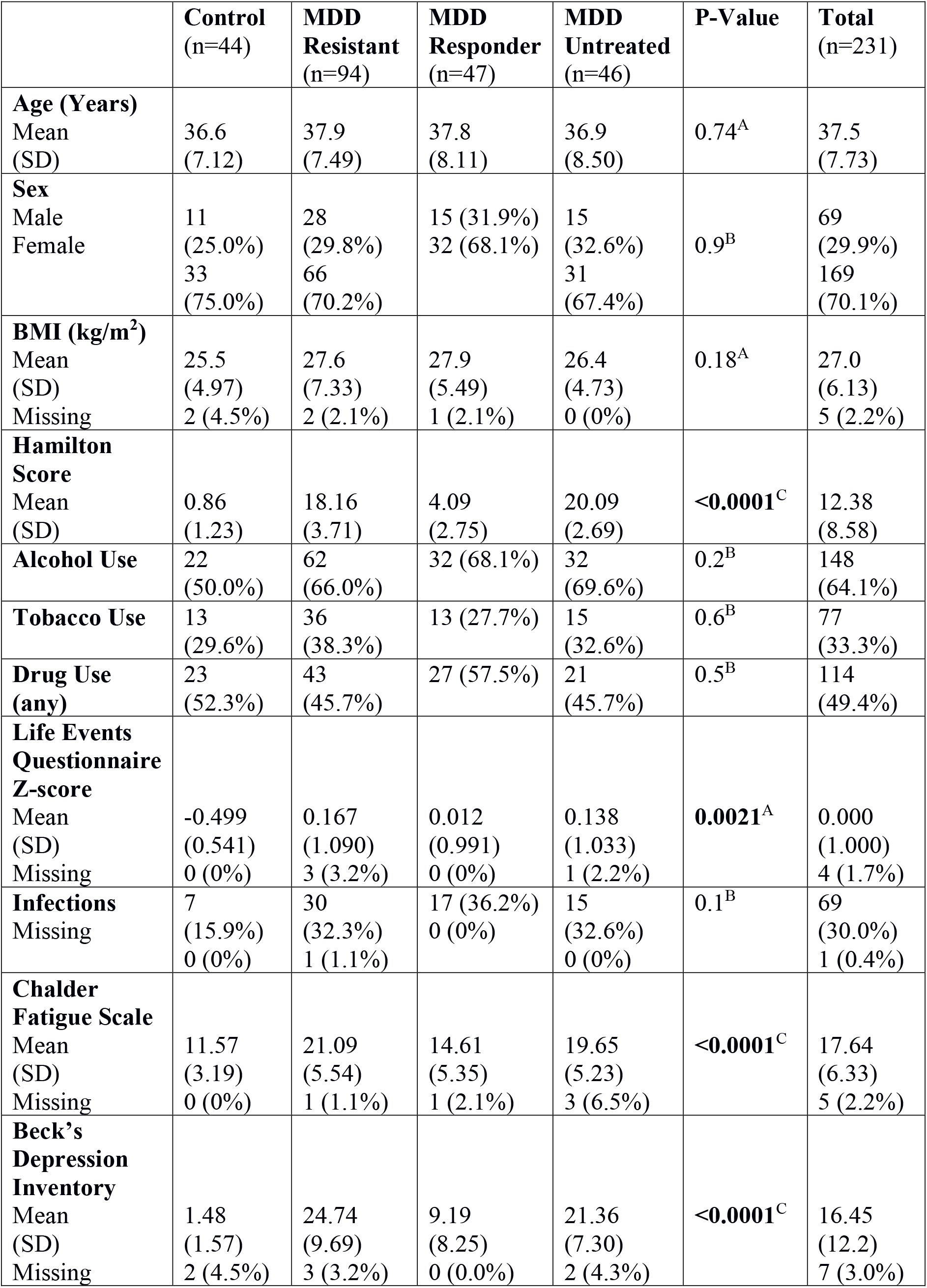
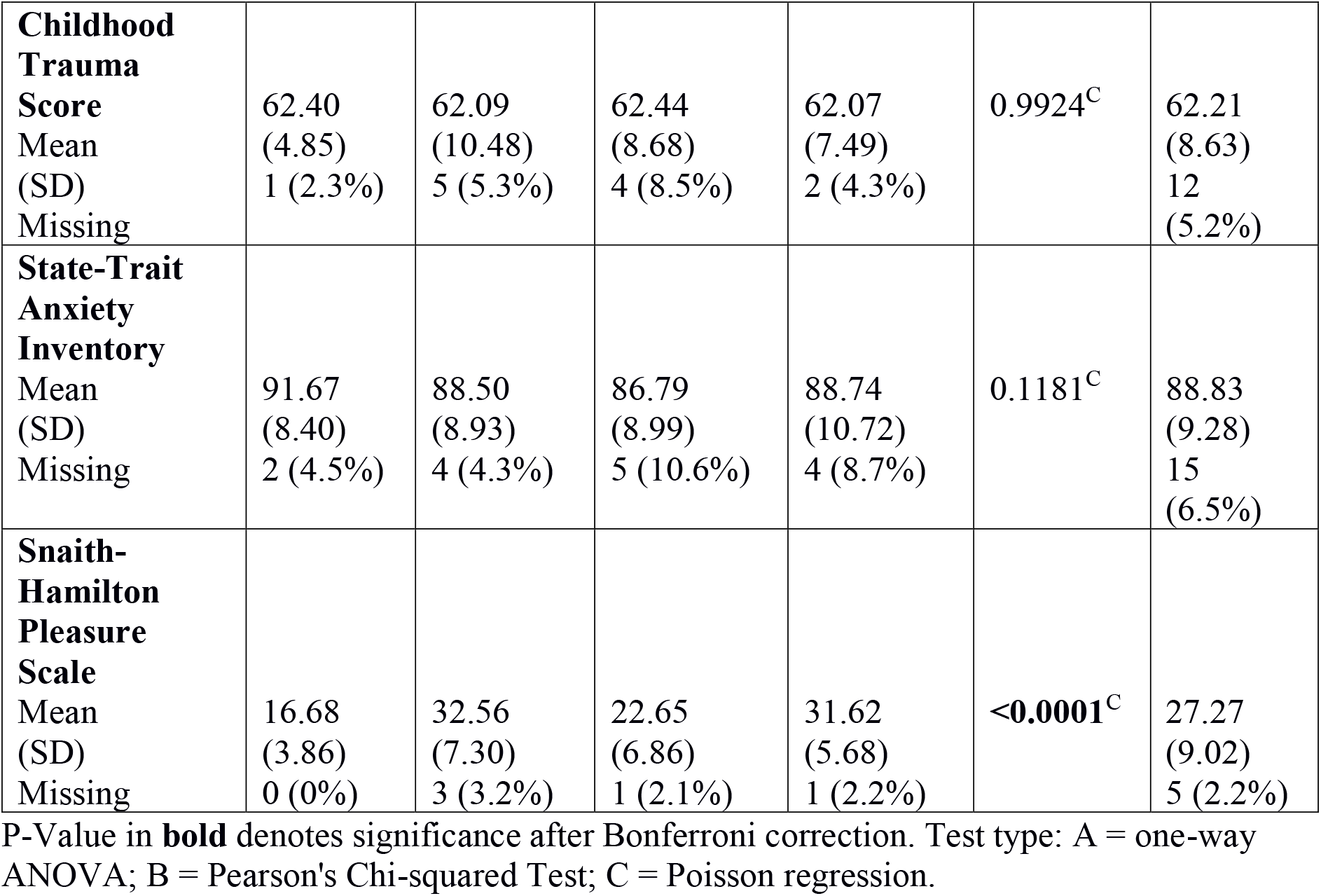
Selection of samples and clinical characteristics.

### Quality control and identification of confounding variables

The NIMA samples were deeply sequenced and aligned to the human genome, exhibiting a high per sample alignment and feature counts rate (> 85% alignment, median of 40 million exonic reads, per sample). Deconvolution analysis^12^ showed the distributions of cell types to be consistent between the samples and typical of PBMCs (**Figure 1A**). Deseq2 Differential expression analysis identified fifteen potentially confounding clinical covariates (each with > 5 significant genes each at adjusted p < 0.01) from a panel of 87 (**Figure 1B**), with Age, Gender and BMI showing the strongest effects by an order of magnitude (1,244, 625 and 203 significant genes respectively). The expression profiles for the Age, Gender and BMI associated genes were consistent across all samples (**Figure 1C**) and the most differential genes (**Figure 1D**) were consistent with the relevant biology (e.g. the most significant gender related genes were UTX and HYA which are X and Y linked^19, 20^). We therefore concluded firstly that the data was of a high quality both technically and experimentally, and secondly that, given the size of the observed effect in the primary data, it was appropriate to control for the fifteen confounding clinical covariates in the downstream analysis.

### There is no robust evidence for a differential expression signature between HC and MDD in PBMCs

We used differential expression analysis to characterise any differences between HC and each of the MDD groups (MDD, treatment-resistant, treatment-responsive and untreated), using an adjusted p cut-off of < 0.01, and including all 15 confounding clinical covariates plus batch as interaction terms. One significantly different gene was evident between HC and MDD (**HIST1H2AE**, adjusted p = 0.008) and none between HC and MDD responders, MDD resistant or MDD untreated (**Supplementary Table 1-4**). We additionally tried reduced differential models – without BMI, with Age, BMI and Gender only and with Batch only, however it made no meaningful difference to these results. Observing only one significant gene suggested that either 1) the adjusted p-value threshold was too strict, or 2) the adjusted p-value threshold was reasonable, and we were observing type I error at HIST1H2AE. When we viewed the per sample expression at HIST 1H2AE (**Figure 2A**) it showed the difference in expression between HC and the MDD groups to be highly subtle. This was also true for the two genes of lowest p-value (non-significant) for each of the four comparisons (**Figure 2A-D**).

**Figure 2.**
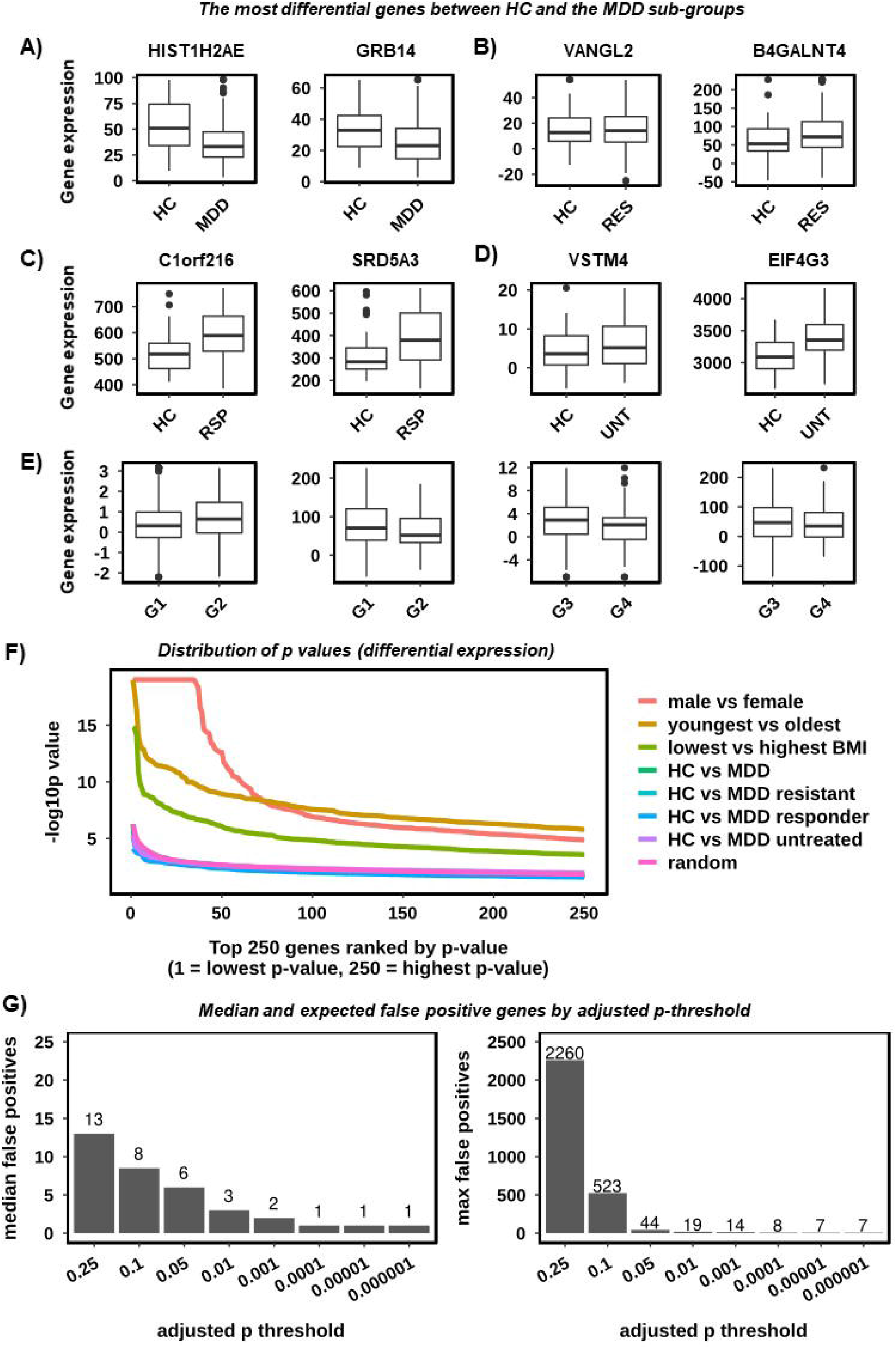
There is no evidence for a classical differential expression signature between HC and MDD in PBMCs. **A)** Gene expression boxplots highlighting the most significantly different genes between HC and MDD. Sample groups are shown on the x-axis and gene expression values (DESeq2 normalised counts) on the y-axis. B) As A) however for HC vs the MDD treatment-resistant group. **C)** As A) however for HC vs the MDD treatment-responsive group. **D)** As A) however for HC vs the MDD untreated group. **E)** As A) however for the two most significant genes from each of two comparisons of randomised cases and controls. Randomised groups are labelled G1-G4. **F)** Distribution of differential expression p-values highlighting the consistency between HC vs MDD and randomised cases and controls. The 250 most significant genes for each comparison are shown on the x-axis (ranked from lowest to highest) and the p value (as -log10) on the y-axis. Lines are given for the three confounding variables Gender (‘male vs female’), Age (‘youngest vs oldest’), BMI (‘lowest vs highest’), HC vs the four MDD types (MDD, MDD treatment-resistant, MDD treatment-responsive and MDD untreated), and for the average of 50 comparisons of randomised cases and controls (‘random’). **G)** Bar charts highlighting the number of differentially expressed genes that were expected to be false positives by adjusted p threshold, based on 50 iterations of randomised cases and controls. The adjusted p threshold is given on the x-axis and the median (left) and maximum (right) number of expected false positives on the y-axis.

**Figure 2E** highlights the two most significant genes from each of two comparisons of randomised cases and controls. Randomised groups are labelled G1-G4. At the 250 most highly significant genes for each comparison the distributions of p-values were almost identical to that of randomised cases and controls (**Figure 2F**). This was in stark contrast to age, gender and BMI. These observations suggested that relaxing the adjusted p-threshold would not increase the number of true positives. We next estimated the number of false positives expected in this dataset at a range of adjusted p thresholds by generating 50 differential expression comparisons using randomised cases and controls and taking the median and maximum numbers of significant genes (**Figure 2G**). The results showed that we would expect on average three false positives at adjusted p < 0.01, suggesting that it was not unlikely for HIST 1H2AE to be false positive in this case. Though it is difficult to prove a negative outright, the balance of probabilities suggest that the data more strongly supported the absence of a HC vs MDD differential expression signature in PBMCs.

### There is no evidence for clusters of highly correlating genes that are altered in MDD compared to HC

We next considered the possibility that a HC vs MDD differential signature in PBMCs could be too subtle to detect using single gene interactions. This could occur for example if it originated from a subset of cells within the population. Several transcriptomic studies have shown^21-24^ that subtle signatures can be reliably detected by collapsing clusters of highly correlating genes into representative metagenes for differential expression analysis. This acts to reduce noise and multi-sample correction stringency at the expense of single gene resolution. To do so we removed genes with low expression (mean > 10, in the Combat corrected data) or with exceptionally high coefficient of variability (standard deviation / mean < 0.15), to reduce the chance that correlations could be driven by technical variability^15^. Next, we generated a gene co-expression matrix from the remaining **5,356** genes and plotted it as a hierarchically clustered heatmap (**Figure 3A**). The heatmap showed clear structure and confirmed the existence of several clusters of highly correlating genes. To identify the correlation clusters, we used the method as described in Le et al^15^ (Supplementary Information). We identified **48** gene clusters with at least 50 genes in each. To validate these clusters, we plotted them as expression heatmaps (**Figure 3B**), which confirmed the highly correlating nature of the genes in each. Next, we set out to determine whether the expression at cluster metagenes differed between HC and MDD. We generated per cluster metagenes and compared the metagene expression for HC samples to MDD samples. We observed no significant difference (p < 0.25, unpaired, two tailed t-test with Benjamini-Hochberg correction) between HC and MDD in any cluster (**data not shown**). Boxplots of the six clusters of lowest p-value (non-significant) highlighted the absence of any convincing biological differences at each cluster (**Figure 3C**). We therefore concluded that there was no evidence for clusters of highly correlating genes that are altered in MDD compared to HC in this dataset.

**Figure 3.**
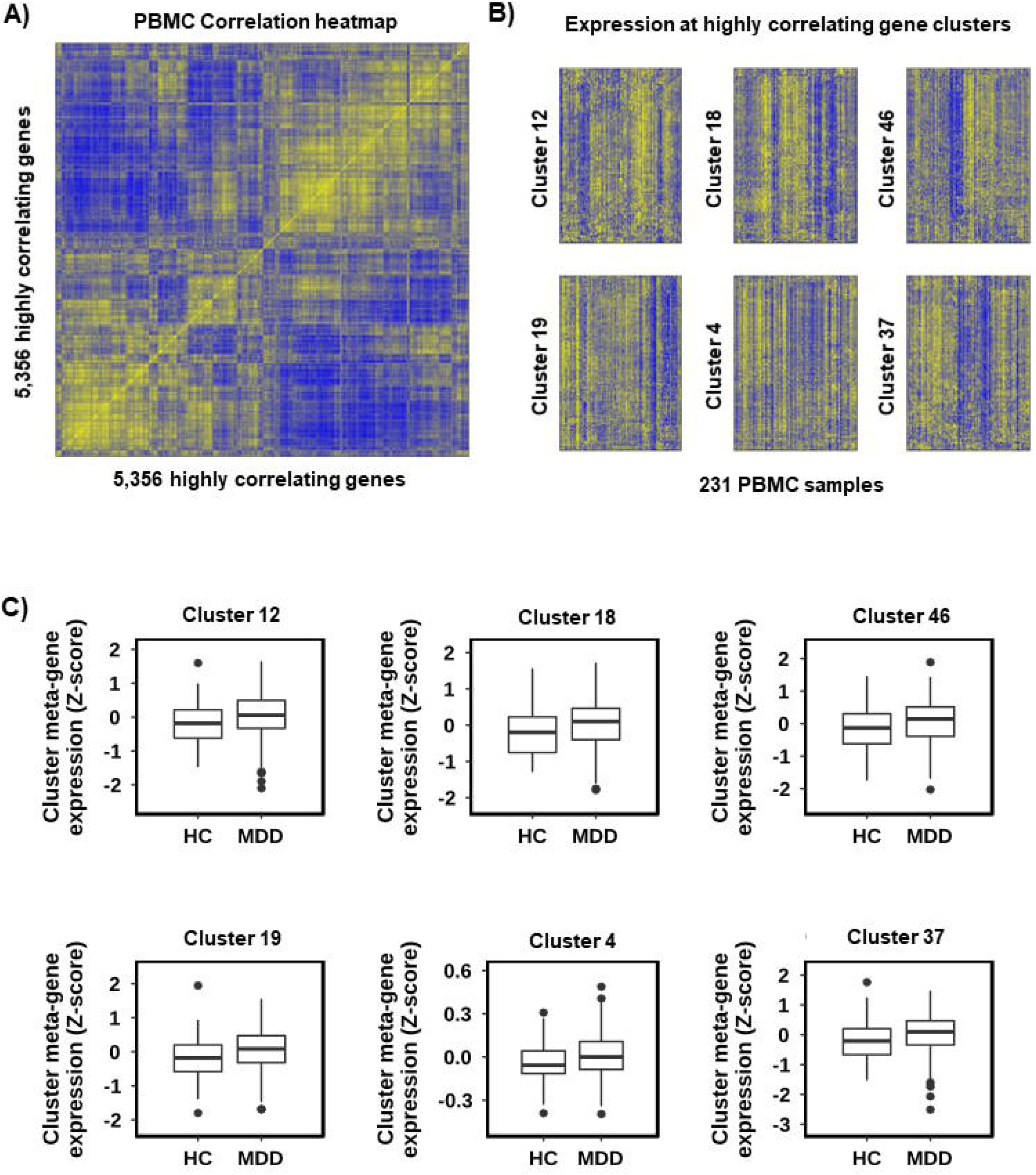
There is no evidence for clusters of highly correlating genes that are altered in MDD compared to HC. **A)** Gene co-expression heatmap highlighting the presence of clusters of highly correlating genes in PBMC data. The x and y-axis show the 5,356 highly correlating genes. The colour intensity indicates the spearman correlation value between two given genes with blue as low and yellow as high. To highlight the presence of co-expression clusters the heatmap has been hierarchically clustered on both axes using Spearman distances, with UPMGA agglomeration and mean reordering. **B)** Gene expression heatmaps for six gene co-expression clusters, highlighting the consistency between the expression pattern of all genes within a cluster across all 231 samples. Samples are given by column and cluster genes by row. Colour intensity indicated row scaled (z-score) gene expression, with blue as low and yellow as high. **C)** Gene expression boxplots for the six clusters with the lowest p-value (T-test) for HC vs MDD. Showing sample group on the x-axis and the cluster metagene expression (mean z-score) on the y-axis. All clusters are non-significant with adjusted p > 0.25.

### False positive genes were not random in PBMC data

We observed in our 50 differential comparisons involving randomised cases and controls that the most significant genes included genes of immune function (such as TNF and IFIT2) more frequently than we expected. This raised the possibility that false positives genes might preferentially be immune genes when looking at PBMCs.

To test this hypothesis, for each gene we took the mean p-value across the fifty randomised comparisons, then selected the 50 most highly significant genes by mean p-value. We ran over representation analysis on the genes (using DAVID with GO biological processes) and found nine significantly enriched (< 5% FDR) gene ontologies (**Table 2**). All were immune related with the top three being “response to virus”, “type I interferon signaling pathway” and “cellular response to interleukin-1” and included the genes IFIT1, IFIT2, IFIT3 and CCL8. This suggested that false positives are not random in these data and show a significant bias towards immune functions. This further supported that it would not be reasonable to relax the adjusted p threshold when comparing HC to MDD, as it would likely introduce an erroneous immune signal that could be confused for bona-fide.

**Table 2.** Enriched Gene ontologies (FDR < 5%) the 154 genes that were consistently significant using randomised cases and controls in PBMC.

| ID | Term | Term Size | Overlap | Enrichment | FDR |
| --- | --- | --- | --- | --- | --- |
| GO:0070098 | chemokine-mediated signaling pathway | 71 | 9 | 14.99 | 0.00022 |
| R-HSA-380108 | chemokine receptors bind chemokines | 59 | 8 | 13.67 | 0.00188 |
| GO:0006954 | inflammatory response | 379 | 13 | 4.06 | 0.13608 |
| GO:0030593 | neutrophil chemotaxis | 66 | 6 | 10.75 | 0.36564 |
| hsa04060 | cytokine-cytokine receptor interaction | 243 | 11 | 3.94 | 0.45579 |
| GO:0051607 | defense response to virus | 165 | 8 | 5.73 | 0.77463 |
| GO:0006935 | chemotaxis | 122 | 7 | 6.79 | 0.90809 |
| hsa04062 | chemokine signaling pathway | 186 | 9 | 4.21 | 1.35807 |
| GO:0048247 | lymphocyte chemotaxis | 28 | 4 | 16.89 | 2.55814 |
| GO:0060337 | type I interferon signaling pathway | 64 | 5 | 9.24 | 3.2096 |
| GO:0060326 | cell chemotaxis | 65 | 5 | 9.10 | 3.39502 |
| GO:0009615 | response to virus | 110 | 6 | 6.45 | 3.67281 |
| GO:0043491 | protein kinase B signaling | 33 | 4 | 14.33 | 4.10142 |
| GO:0071347 | cellular response to interleukin-1 | 71 | 5 | 8.33 | 4.65869 |
| GO:0060009 | sertoli cell development | 10 | 3 | 35.48 | 4.68831 |
| R-HSA-909733 | interferon alpha/beta signaling | 67 | 5 | 7.52 | 4.90525 |

### Relative to patient age biological age is significantly greater in MDD patients than HC

To explore whether MDD patients showed increased biological ageing compared to HC, we estimated the biological age of each sample by taking the mean expression value (z-score) across all the age-related genes (see Methods and Supplementary Methods for full details) and plotted it against chronological age (**Figure 4A**). As expected, we observed a strong positive and significant linear correlation between biological and chronological age (Spearman Correlation Coefficient (SCC) = 0.72, p < 0.01). To determine whether MDD or HC patients showed altered biological ageing (relative to chronological age) we performed a linear regression using the model biological age ~ chronological age (**Figure 4A**). Next, we counted the number of HC or MDD patients above or below the regression line and found a subtle (HC – 26 below (59%), 18 above (41%), MDD – 92 below (49%), 95 above (51%)) but significant difference (p <0.01, Fisher’s exact test). To illustrate the difference in distribution, we used the residuals – i.e. the distance along the y-axis of each dot from the regression line (**Figure 4B**). Finally, to validate the result we replicated the analysis using the GSK-HiTDiP MDD^16^ whole blood microarray data. The results were comparable to PBMCs (**Figure 4A-B**), with the MDD patients showing significantly elevated biological ageing relative to chronological ageing (HC – 35 below (61%), 22 above (39%), MDD – 48 below (42%), 65 above (58%)), p<0.01, Fisher’s exact test).

**Figure 4.**
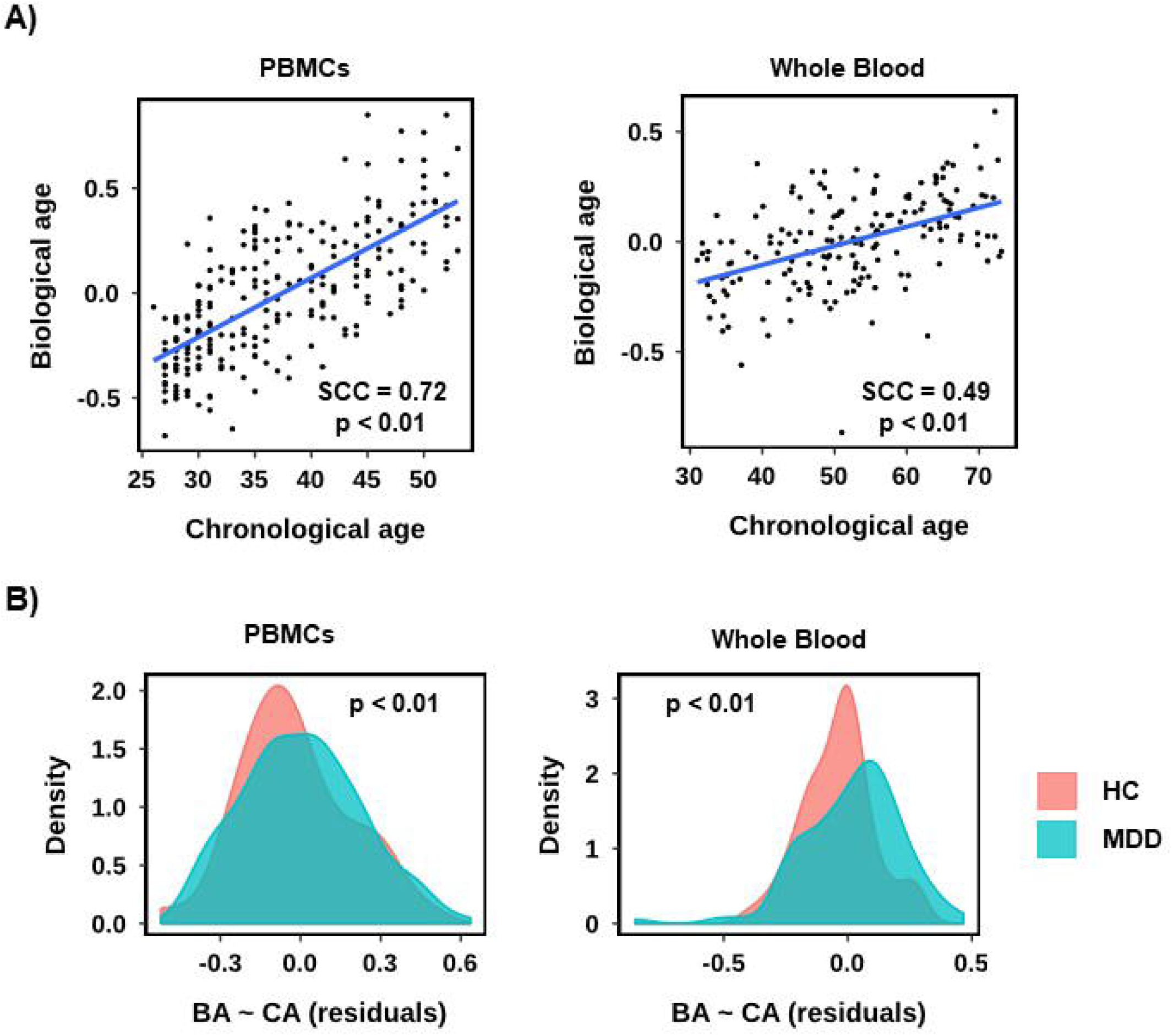
Relative to patient age biological age is greater in MDD patients than in HC. **A)** Scatterplots for PBMC RNA-seq data (left) and whole blood expression microarray data (right), showing the correlation between chronological age (x-axis) and biological age (y-axis) as defined by the mean expression z-score across all age-related genes, per sample. A linear regression line, alongside the Spearman Correlation Coefficient (SCC) and associated p-value is shown. **B)** Density plots of the residuals from the linear regressions in A). A positive residual indicates a sample above the regression line and negative below.

## Discussion

In this large, well-controlled and deeply sequenced data-set, we find no evidence for a differential expression signature in PBMCs between HC and MDD – as a whole or in the subtypes described; nor is there evidence for clusters of highly correlating genes that are altered in MDD compared to HC. We also found that biological age relative to chronological age is significantly greater in MDD patients than in HC.

Our differential analysis showed only one gene to differ significantly (adjusted p < 0.01) between HC and MDD and none between HC and MDD sub-groups. Further investigation concluded that, given the concurrence between the distribution of p values for random samples and the HC and MDD group comparisons, the very low difference in expression between HC and MDD groups at these genes, and the numbers of expected false positives at this adjusted p-threshold, this was most likely a false positive, unlikely to be biologically meaningful, and that there was no justification for relaxing the p value threshold in this data. To test whether any HC vs MDD signature might be too subtle to detect at the single gene level, we generated 48 gene co-expression clusters and compared the metagenes between HC and MDD. We observed no significant differences in any cluster, or any convincing biological differences. We therefore concluded that the data more strongly supported the absence of a HC vs MDD differential expression signature in PBMCs.

In addition, when we randomised cases and controls fifty times and performed over representation analysis, we found the most significant false positives to not be random but to have a significant immune phenotype, including “response to virus” and “type I interferon signalling pathway”. This further justified not relaxing the adjusted p threshold in this data, as doing so would likely introduce an erroneous immune signature that could be interpreted as bona fide.

These results are, in many ways, comparable to previous transcriptomic studies in whole blood which also found no signature at adjusted p < 0.05 using larger sample numbers^2^. One strength of our approach is that we control for age, gender and BMI in our sample selection. In our opinion, we could not justify relaxing our adjusted p threshold. However, other studies identified signatures at adjusted p values ranging from p < 0.1 to p < 0.25.

A further strength of our study is that we present the per sample expression values for all genes of interest. We would argue that as other data^2,26^ presented signatures that were detectable only at adjusted p > 0.05 using around 1,000 samples each, these signatures are likely to be subtle. However, the omission of per sample expression data at the genes of interest, makes it difficult to establish how subtle and so it is difficult to form a robust opinion of how biologically meaningful these expression differences are.

As mentioned in the introduction, evidence for an inflammatory protein signature in MDD is substantial. This is particularly the case for the proinflammatory cytokine IL-6, with several meta-analyses confirming this. There is also a longitudinal association between MDD and IL-6^27^, yet the tissue source of cytokines remains unclear. Our data strongly suggest that in this sample of MDD, the source of cytokines is unlikely to be PBMCs. Reflecting on other potential sources; **neutrophils** are increasingly seen as important for fine regulation of the immune-inflammatory response, outnumbering PBMCs by one or two orders of magnitude^28^. Neutrophils produce a large variety of chemokines and cytokines upon stimulation and can differentially switch phenotypes, displaying distinct subpopulations in different microenvironments^29^. If neutrophils confer the cytokine signature, it would be expected that gene expression studies of whole blood would capture their contribution. Another potential cell source are **endothelial cells**. These are ubiquitous in both brain and periphery. Recently, Blank et al demonstrated a specific role in relation to aspects of depression-relevant behaviour in mice by showing that downstream signalling of brain endothelial cells induces fatigue and cognitive impairment via impaired neurotransmission in the hippocampus^30^. However, assessing the individual contribution of endothelial cells in humans would be technically very challenging. Nevertheless, considering findings presented in a recent GWAS of MDD, it is important to consider that peripheral tissues may have less of an overall contribution than the brain. Wray et al integrated their GWAS data with functional genomic data, comparing their findings with bulk tissue RNAseq from genotype tissue expression (GTEx)^1^. Here only brain tissue showed enrichment, with the areas showing the most significant enrichment being cortical. This was in contradistinction to other tissue types including whole blood.

The issue of body mass in MDD is complex. Wray et al found significant positive genetic correlations with body mass^1^ and Mendelian randomization (MR) analysis was consistent with BMI being causal or correlated with causal risk factors for depression. Also, negative MR results provide important evidence of no direct causal relationship between MDD and *subsequent* changes in BMI. Adipose tissue actively secretes cytokines and obesity is itself associated with changes in the secretome of adipocytes leading to increased production of proinflammatory cytokines^31^. This raises the possibility that **adipocytes** may be a potential source of inflammatory cytokines acting as a tissue “reservoir”. Careful consideration should be applied when deciding whether BMI should be treated as a confounding variable in MDD or incorporated as part of disease pathogenesis.

We demonstrated that MDD samples showed significantly elevated biological age compared to HC. Although significant, the effect was relatively subtle, comparable to that identified in CpG methylation data. Diniz et al (2019) found MDD exhibited greater molecular senescence in young and middle-aged adults by examining the impact of MDD on the senescence associated secretory phenotype (SASP), a dynamic secretory molecular pathway indicative of cellular senescence^4^. More severe episodes of depression present with higher SASP indices and a significant interaction between current MDD episode and overweight, thus comorbid current MDD plus being overweight had the highest SASP index. While we have not correlated with direct measures of senescence such as SASP indices or epigenetic markers, we would argue that our finding is consistent with the literature and points to a potentially interesting biology.

The strengths of this study lie in the high-quality RNA and large clinical dataset, sequenced to an average depth of > 54.5 million reads, which aligned with >70% of the reads mapping to exons. Thus, a deeply sequenced, well-controlled clinical sample. The limitations of this study relate to heterogeneity inherent in MDD. Within our study, there was also some heterogeneity within the assessing of prior medications as this was done using retrospective self-reporting, albeit based on a comprehensive structured instrument completed by an interviewer.

## Conclusion

This study was a detailed and careful examination of the transcriptomic signal in PBMCs in MDD and HCs. The lack of a significant differentiating signal between MDD and HCs was confirmed by the randomisation of the cases and controls. There was, however, evidence of elevated biological ageing relative to patient age in MDD vs HC. Future work should endeavour to expand clinical sample sizes, reduce MDD heterogeneity and account for confounds from the outset. Advances in RNA-seq at the level of the single cell may help uncover further, more subtle differences. However, the subtlety of any signature mitigates against convincing use as a diagnostic or predictive biomarker, and tissue enriched data is strongly indicative of brain tissue being the most informative in this regard.

## Data Availability

All raw data are held by the NIMA Consortium and can be provided by the corresponding author on request

## Acknowledgements

The BIODEP study was sponsored by the Cambridgeshire and Peterborough NHS Foundation Trust and the University of Cambridge and funded by a strategic award from the Wellcome Trust (104025) in partnership with Janssen, GlaxoSmithKline, Lundbeck and Pfizer. Recruitment of participants was supported by the National Institute of Health Research (NIHR) Clinical Research Network: Kent, Surrey and Sussex & Eastern. Additional funding was provided by the National Institute for Health Research (NIHR) Biomedical Research Centre at South London and Maudsley NHS Foundation Trust and King’s College London, and by the NIHR Cambridge Biomedical Research Centre (Mental Health). ETB is supported by a Senior Investigator award from the NIHR.

We would like to gratefully thank all study participants, research teams and laboratory staff, without whom this research would not have been possible. All members of the NIMA Consortium at the time of data collection are thanked and acknowledged in the Supplementary NIMA Members List.

Study data were collected and managed using REDCap electronic data capture tools hosted at the University of Cambridge. (Paul A. Harris, Robert Taylor, Robert Thielke, Jonathon Payne, Nathaniel Gonzalez, Jose G. Conde, Research electronic data capture (REDCap) – A metadata-driven methodology and workflow process for providing translational research informatics support, J Biomed Inform. 2009 Apr;42(2):377-81).

## References

1. Wray NR, Ripke S, Mattheisen M, Trzaskowski M, Byrne EM, Abdellaoui A et al. Genome-wide association analyses identify 44 risk variants and refine the genetic architecture of major depression. Nature Genetics 2018; 50(5): 668–681.

2. Mostafavi S, Battle A, Zhu X, Potash JB, Weissman MM, Shi J et al. Type I interferon signaling genes in recurrent major depression: increased expression detected by whole-blood RNA sequencing. Mol Psychiatry 2014; 19(12): 1267–1274.

3. Benayoun BA, Pollina EA, Singh PP, Mahmoudi S, Harel I, Casey KM et al. Remodeling of epigenome and transcriptome landscapes with aging in mice reveals widespread induction of inflammatory responses. Genome Res 2019; 29(4): 697–709.

4. Diniz BS, Reynolds Iii CF, Sibille E, Bot M, Penninx B. Major depression and enhanced molecular senescence abnormalities in young and middle-aged adults. Transl Psychiatry 2019; 9(1): 198.

5. McKim DB, Weber MD, Niraula A, Sawicki CM, Liu X, Jarrett BL et al. Microglial recruitment of IL-1beta-producing monocytes to brain endothelium causes stress-induced anxiety. Mol Psychiatry 2018; 23(6): 1421–1431.

6. Menard C, Pfau ML, Hodes GE, Kana V, Wang VX, Bouchard S et al. Social stress induces neurovascular pathology promoting depression. Nat Neurosci 2017; 20(12): 1752–1760.

7. Garre JM, Silva HM, Lafaille JJ, Yang G. CX3CR1(+) monocytes modulate learning and learning-dependent dendritic spine remodeling via TNF-alpha. Nat Med 2017; 23(6): 714-722.

8. Chamberlain SR, Cavanagh J, de Boer P, Mondelli V, Jones DNC, Drevets WC et al. Treatment-resistant depression and peripheral C-reactive protein. Br J Psychiatry 2019; 214(1): 11–19.

9. Martin M. Cutadapt removes adapter sequences from high-throughput sequencing reads. 2011 2011; 17(1): 3.

10. Dobin A, Davis CA, Schlesinger F, Drenkow J, Zaleski C, Jha S et al. STAR: ultrafast universal RNA-seq aligner. Bioinformatics 2012; 29(1): 15–21.

11. Love MI, Huber W, Anders S. Moderated estimation of fold change and dispersion for RNA-seq data with DESeq2. Genome Biology 2014; 15(12): 550.

12. Chen B, Khodadoust MS, Liu CL, Newman AM, Alizadeh AA. Profiling Tumor Infiltrating Immune Cells with CIBERSORT. Methods Mol Biol 2018; 1711: 243-259.

13. Huang da W, Sherman BT, Lempicki RA. Systematic and integrative analysis of large gene lists using DAVID bioinformatics resources. Nat Protoc 2009; 4(1): 44-57.

14. Huang da W, Sherman BT, Lempicki RA. Bioinformatics enrichment tools: paths toward the comprehensive functional analysis of large gene lists. Nucleic Acids Res 2009; 37(1): 1-13.

15. Le TT, Savitz J, Suzuki H, Misaki M, Teague TK, White BC et al. Identification and replication of RNA-Seq gene network modules associated with depression severity. Transl Psychiatry 2018; 8(1): 180.

16. Leday GGR, Vertes PE, Richardson S, Greene JR, Regan T, Khan S et al. Replicable and Coupled Changes in Innate and Adaptive Immune Gene Expression in Two Case-Control Studies of Blood Microarrays in Major Depressive Disorder. Biol Psychiatry 2018; 83(1): 70–80.

17. Ritchie ME, Phipson B, Wu D, Hu Y, Law CW, Shi W et al. limma powers differential expression analyses for RNA-sequencing and microarray studies. Nucleic Acids Research 2015; 43(7): e47-e47.

18. Li Q, Birkbak NJ, Gyorffy B, Szallasi Z, Eklund AC. Jetset: selecting the optimal microarray probe set to represent a gene. BMC Bioinformatics 2011; 12(1): 474.

19. Jansen R, Batista S, Brooks AI, Tischfield JA, Willemsen G, van Grootheest G et al. Sex differences in the human peripheral blood transcriptome. BMC Genomics 2014; 15(1): 33.

20. Mendelson MM, Marioni RE, Joehanes R, Liu C, Hedman AK, Aslibekyan S et al. Association of Body Mass Index with DNA Methylation and Gene Expression in Blood Cells and Relations to Cardiometabolic Disease: A Mendelian Randomization Approach. PLoS Med 2017; 14(1): e1002215.

21. Chen C, Cheng L, Grennan K, Pibiri F, Zhang C, Badner JA et al. Two gene coexpression modules differentiate psychotics and controls. Mol Psychiatry 2013; 18(12): 1308–1314.

22. Gaiteri C, Ding Y, French B, Tseng GC, Sibille E. Beyond modules and hubs: the potential of gene coexpression networks for investigating molecular mechanisms of complex brain disorders. Genes Brain Behav 2014; 13(1): 13-24.

23. Roy S, Bhattacharyya DK, Kalita JK. Reconstruction of gene co-expression network from microarray data using local expression patterns. BMC Bioinformatics 2014; 15 Suppl 7: S10.

24. Wang X, Dalkic E, Wu M, Chan C. Gene module level analysis: identification to networks and dynamics. Curr Opin Biotechnol 2008; 19(5): 482-491.

25. Han LKM, Aghajani M, Clark SL, Chan RF, Hattab MW, Shabalin AA et al. Epigenetic Aging in Major Depressive Disorder. Am J Psychiatry 2018; 175(8): 774-782.

26. Jansen R, Penninx BWJH, Madar V, Xia K, Milaneschi Y, Hottenga JJ, Hammerschlag AR, Beekman A, van der Wee N, Smit JH, Brooks AI, Tischfield J, Posthuma D, Schoevers R, van Grootheest G, Willemsen G, de Geus EJ, Boomsma DI, Wright FA, Zou F, Sun W, Sullivan PF. Gene expression in major depressive disorder. Molecular Psychiatry 2016; 21: 339–347.

27. Lamers F, Milaneschi Y, Smit JH, Schoevers RA, Wittenberg G, Penninx BWJH. Longitudinal Association Between Depression and Inflammatory Markers: Results From the Netherlands Study of Depression and Anxiety. Biological Psychiatry 2019; 85(10): 829-837.

28. Tecchio C, Micheletti A, Cassatella MA. Neutrophil-derived cytokines: facts beyond expression. Front Immunol 2014; 5: 508-508.

29. Rosales C. Neutrophil: A Cell with Many Roles in Inflammation or Several Cell Types? Front Physiol 2018; 9: 113-113.

30. Blank T, Detje CN, Spiess A, Hagemeyer N, Brendecke SM, Wolfart J et al. Brain Endothelial- and Epithelial-Specific Interferon Receptor Chain 1 Drives Virus-Induced Sickness Behavior and Cognitive Impairment. Immunity 2016; 44(4): 901-912.

31. Fuster JJ, Ouchi N, Gokce N, Walsh K. Obesity-Induced Changes in Adipose Tissue Microenvironment and Their Impact on Cardiovascular Disease. Circ Res 2016; 118(11): 1786-1807.

